# Health Surveillance and Epidemiological Profile of Severe Acute Respiratory Syndrome due to COVID-19 in Brazil (2020-2024)

**DOI:** 10.1101/2025.04.27.25326529

**Authors:** Silvio Alencar Cândido-Sobrinho, José Quirino da Silva-Filho, Francisco de Sousa Júnior, Vânia Angélica Feitosa Viana, Aldo Ângelo Moreira Lima

**Author notes:** **Corresponding Author**: Silvio Alencar Cândido-Sobrinho, Post-graduate Program in Medical Sciences, Faculty of Medicine, Federal University of Ceará, Rua Prof. Costa Mendes, 1608 - 4^th^ floor, Fortaleza, Ceará, Brazil.

## Abstract

**WHAT WAS ALREADY KNOWN?:**

- **Variants influence COVID-19 severity:** Variants such as Delta and Gamma were already associated with increased transmissibility and lethality.
- **Vaccination has a protective role against severe forms of the disease:** Previous data showed reduced mortality with vaccines, especially after completing the primary series.
- **Regional inequalities impact outcomes:** Regions with lower healthcare infrastructure, especially in the North and Northeast, face higher fatality rates.

**WHAT IS NEW?:**

- **Three major waves of SARS due to COVID-19, with marked differences by region and variant:** The second wave was the deadliest, and Omicron, though less pathogenic, had widespread circulation and three distinct peaks.
- **Direct impact of sociodemographic and economic factors on outcomes:** Lethality was higher among illiterate individuals, Black and Brown populations, and rural residents.
- **Robust effect of booster vaccination on survival:** Individuals with a first or second booster were up to 60% less likely to die and had four times higher survival at 40 days compared to unvaccinated individuals.

The COVID-19 crisis exposed deep-rooted inequalities in Brazil, such as the concentration of healthcare resources in metropolitan areas. Despite the availability of effective vaccines, full immunization coverage was not achieved, allowing cases to progress to severe acute respiratory syndrome (SARS). This study aimed to analyse the progression of mild, moderate, and severe cases between January 2020 and December 2024, covering a five-year period, taking into account regional, social, and variant-related differences, vaccination, and factors associated with SARS due to COVID-19 in Brazil. A total of 47,547,814 influenza-like illness (mild and moderate) cases, 2,127,427 SARS cases due to COVID-19, 533,966,291 vaccination records, and data on circulating variants were analysed. Among the SARS cases, 1,171,801 were confirmed by PCR; 777,672 patients recovered, and 394,129 died, resulting in a case fatality rate of 33.63%. Brazil experienced three major waves of SARS due to COVID-19, with the second wave being the deadliest across all regions. The Gamma and Omicron variants were the most persistent and impactful. The transition between variants influenced the regional dynamics of the pandemic, although little variation was observed in the proportion of circulating variants across regions. The study highlights the importance of continuous monitoring, genomic surveillance, and vaccination coverage to anticipate and mitigate future pandemic waves.

## INTRODUCTION

The emergence of the Influenza A (H1N1) virus in 2009 triggered a rapid global spread, reigniting concerns about the potential of etiological agents to cause pandemics. In response to this emergency, Brazil’s Ministry of Health established the Severe Acute Respiratory Syndrome (SARS) Surveillance Group, focused on the prevention and control of emerging respiratory diseases^1^.

Years later, the health crisis caused by COVID-19 would expose deep structural inequalities in the country: the concentration of Intensive Care Units (ICUs) in metropolitan areas^2^, shortage of human resources, medications, and hospital supplies^3,4^, vaccine hesitancy^5^ and the emergence of new SARS-CoV-2 variants^6^. Despite the availability of effective vaccines, full immunization was not universal, allowing COVID-19 to progress to severe cases as SARS^7^. Brazil became the epicentre of the pandemic in 2021, largely due to its vast, heterogeneous territory and political scenario, which directly influenced decision-making processes during the health crisis^8^. Social inequalities further exacerbated this scenario, both through the concentration of healthcare resources and economic vulnerability, which forced a significant portion of the population to continue working despite recommendations for social distancing^9^.

In this context, it is essential to understand how COVID-19 progresses to SARS among the Brazilian population by identifying risk groups and factors associated with unfavourable outcomes. Additionally, analysing the evolution of cases and deaths, crisis management strategies, the emergence of new variants, and the vaccination campaign is crucial to identifying gaps in the pandemic response and informing future public health policies. Thus, this study aims to describe the dynamics of COVID-19 in Brazil over five years (2020-2024), considering regional and social differences, the impact of circulating variants, vaccination, and factors associated with progression to SARS.

## METHODS

### Research Ethics

This study used aggregated data without the possibility of individual identification, made available in public repositories, and was therefore exempt from evaluation by a Research Ethics Committee according to the Brazilian National Health Council Resolution No. 510/2016^10^.

### Study Design

This is an observational study of the retrospective cohort^11^, based on secondary data extracted from national health surveillance systems, encompassing mild, moderate, and severe cases of COVID-19 between 2020 and 2024 in Brazil.

### Data Source

Data covering the period from January 1, 2020, to December 31, 2024, were retrieved as follows: mild and moderate cases were obtained from the ‘Influenza-like Illness Notifications Database’ (referred as ‘ILI’); severe cases were obtained from the ‘Severe Acute Respiratory Syndrome Database - including COVID-19 data’ (referred as ‘SARS’); and Vaccination data were obtained from the ‘National COVID-19 Vaccination Campaign Database’, all retrieved from openDataSUS repositories^12^. Information on circulating variants of SARS-CoV-2 in Brazil during the study period was retrieved from the Fiocruz Genomic Network repositories^13^. Human Development Index (HDI) 2010 data by municipality were obtained from the United Nations Development Programme^14^. Population data were retrieved from the IBGE Automatic Recovery System using *sidrar*^15^.

### Inclusion and Exclusion Criteria

For the ILI database (mild and moderate cases), all confirmed COVID-19 cases were included regardless of age, sex, or region, with laboratory-confirmed etiological agent, and whose outcome was cure, death, or home treatment. Records were excluded if it was not possible to identify the state of residence, notification date, or etiological agent (unspecified SARS or suspected cases).

Regarding the SARS database (severe cases), individuals confirmed for SARS-CoV-2 by PCR were included if they had a final classification of ‘Cure’ or ‘Death’, with an outcome duration between 0 and 38 days (IQR = 15 [10-20] days), and presented vaccination records with sequential applications (necessarily following the order: ‘None’, ‘1^st^ Dose’, ‘2^nd^ Dose’, ‘1^st^ Booster’, and ‘2^nd^ Booster’), with manufacturer information available and with a minimum period of 7 days after application. Excluded were individuals without PCR confirmation, those who died from other causes, cases without outcome information (ignored cases), and cases with outlier length of stay or inconsistencies in vaccination data.

Regarding the National COVID-19 Vaccination Campaign data, individuals of any sex or region who received the 1^st^ Dose, 2^nd^ Dose, 1^st^ Booster, or 2^nd^ Booster were included. Individuals for whom age or state of residence could not be identified, as well as records of doses related to exceptional cases or new vaccination schedules were excluded.

### Data Processing and Statistical Analysis

The data were processed using AWK, bash, and R^16^ programming languages, with the *tidyverse* libraries used for data processing and string detection^17^. Graphs were generated with *ggplot2*^18^, *ggpubr*^19^, *survminer*^20^ and *forestmodel*^21^ libraries. Contingency tables were created using *tableone*^22^.

Variables were expressed as mean ± standard deviation or median [IQR1-IQR3]. Adjusted incidence and mortality rates were calculated based on data from the 2022 Census. Differences in proportions were assessed using the chi-square test (χ²), and comparisons of case fatality rates (CFR) between groups were performed using the Z-Test for the comparison of multiple two-sided proportions with p-value adjustment by the Bonferroni method using *rstatix*^23^ and *rcompanion*^24^. Analysis of Variance (ANOVA) was performed using the *stats* library, and Tukey’s test was conducted using *agricolae*^25^ library.

Odds Ratios (OR) were reported as OR [95% CI lower - 95% CI upper], estimated through Generalized Linear Models (GLMs). Only complete cases (n=351,051) were considered for this step. Univariate models were fitted including 78 demographic variables (Region, Geographic Zone, Age Group, Sex, Pregnancy Status, 2010 HDI, Ethnicity, and Educational Level), clinical signs and symptoms (General: fever, chills, malaise, sweating, asthenia, prostration, severe fatigue, loss of appetite, hyporexia, insomnia; Respiratory: cough, coryza, bronchitis, sneezing, sore throat, wheezing, tachypnea, respiratory distress, chest pain, cyanosis, hemoptysis, SpO2 saturation < 95%; Neurological: headache, confusion, seizure; Musculoskeletal: myalgia, arthralgia, back pain; Gastrointestinal: nausea, vomiting, abdominal pain, diarrhea; Other: tachycardia, edema, conjunctivitis, sepsis, loss of smell and taste) and risk factors (asthma, cardiopathy, pneumopathy, chronic kidney disease, immunosuppression, chronic liver disease, neurological disorder, obesity, puerperium, Down syndrome, diabetes, Alzheimer’s disease, pancreatitis, alcoholism, anemia, COPD, stroke, hypertension, emphysema, hypothyroidism, arthritis, ascites, lymphoma, lupus, and cancer). Variables with p<0.05 and OR>1.0 were subsequently selected for the multivariate modelling stage, totalling 31 variables. The final model was adjusted using forward-backward stepwise regression with the *MASS*^26^ library.

For modelling purposes, individuals were categorized according to their vaccination status into three groups: those who received no vaccination, those who received the first and/or second dose (‘1^st^/2^nd^ Dose’), and those who received the first and/or second booster (‘1^st^/2^nd^ Booster’). Additionally, the number of comorbidities/risk factors per individual was calculated, including diabetes, neurological disorder, chronic kidney disease, chronic lung disease, immunosuppression, and obesity. These risk factors were selected based on a GLM model adjusted by stepwise selection. Individuals were then classified into three groups according to the total number of reported comorbidities/risk factors: ‘0’ (no comorbidities/risk factors), ‘1-2’(one or two comorbidities/risk factors), and ‘3+’ (three or more comorbidities/risk factors).

Finally, the effect of vaccination status combined with the number of comorbidities/risk factors on median survival time was evaluated through multivariate adjustment of Kaplan-Meier survival curves in the population of severe cases. Comparisons between survival curves were validated using the log-rank test, employing the *survival*^27^ and *survminer*^20^ libraries.

## RESULTS

During the study period (01/01/2020 to 12/31/2024), a total of 47,547,814 cases of Influenza-like Illness (ILI) were reported, of which 25,082,124 were laboratory-confirmed (Figure S1). Simultaneously, 4,029,767 cases of Severe Acute Respiratory Syndrome (SARS) were recorded, with 2,127,427 cases identified as being caused by SARS-CoV-2, and approximately half of these (1,171,801) confirmed by PCR methodology (Figure S1). Regarding the immunization coverage during the period, 184,375,251 1^st^ Doses (90.78% coverage), 166,442,064 2^nd^ Doses (81.95% coverage), 139,285,497 1^st^ Booster doses (68.58% coverage), and 43,863,479 2^nd^ Booster doses (21.59% coverage) were administered. For this study, the 1st+2nd+3rd dose schedules, revaccination, and additional doses were excluded. These regimens are intended for immunocompromised individuals, those undergoing stem cell transplantation, cases of vaccination errors (such as leakage or inadvertent vaccination), and other specific reasons.

Concerning missing data in SARS records for PCR-confirmed cases, 108 individuals had no information regarding sex at birth, 231,100 (19.7%) lacked information on ethnicity, 744,232 (63.5%) lacked education data, and 132,005 (11.3%) had no information regarding their geographic zone of residence. Additionally, a significant difference was observed between the notification date and the data entry date. The largest average delay was recorded in the North Region, with 16.63 ± 15.06ᵃ days, followed by the Southeast Region with 15.32 ± 13.43ᵇ days. No significant difference was found between the Central-West and Northeast Regions, with average delays of 12.58 ± 11.21ᶜ and 12.58 ± 14.01ᶜ days, respectively. The South Region had the fastest reporting time, with an average delay of 12.22 ± 10.25ᵈ days (ANOVA, *post-hoc* Tukey, p<0.05).

The beginning of the pandemic was marked by a surge in cases and deaths, with well-defined curves observed for ILI-related deaths and new cases and deaths from SARS (Figure 1). incidence and mortality rates for SARS due to COVID-19 revealed three distinct waves, with pronounced peaks and regional variations (Figures S2-S3).

**Figure 1.**
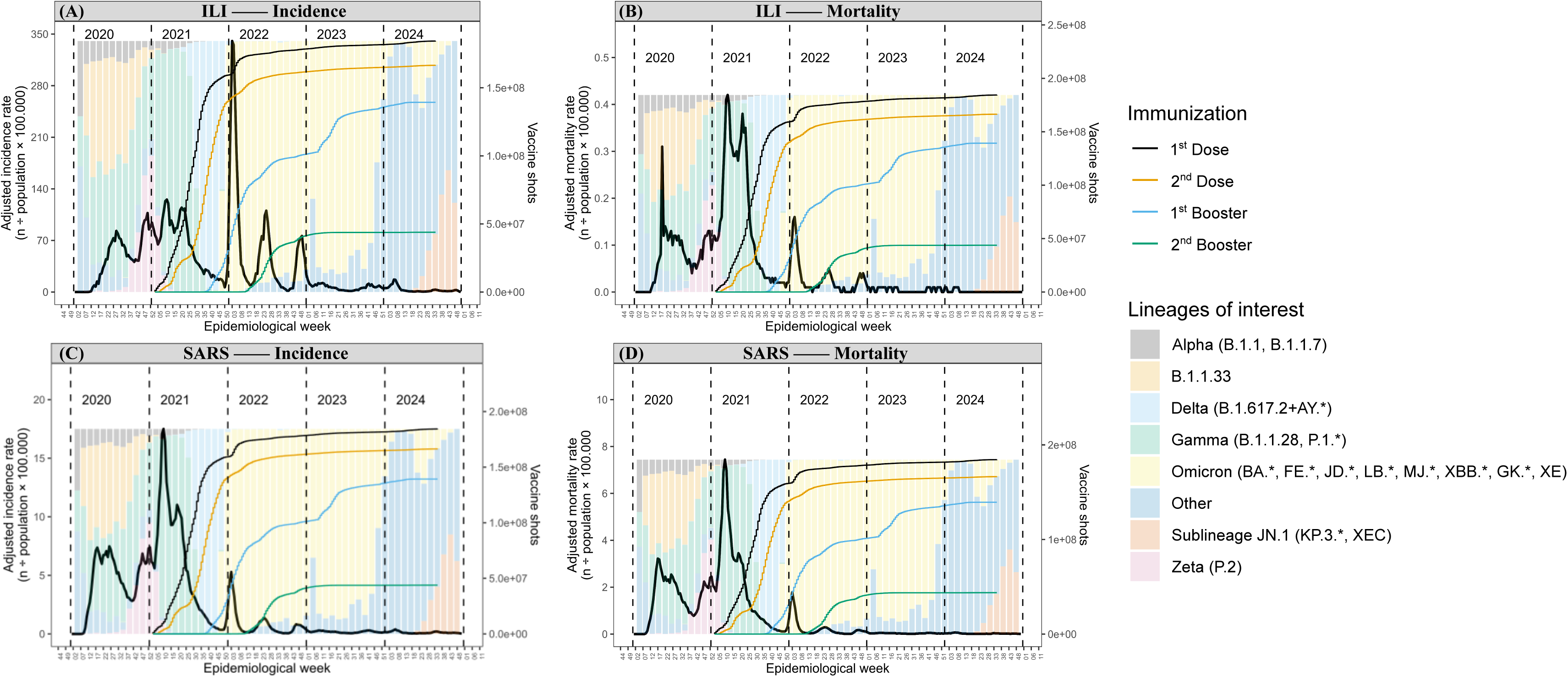
Temporal dynamics of incidence and mortality rates adjusted for population (grey line, left y-axis) by epidemiological week (x-axis), levels of immunization by vaccination stage (lines with right y-axis), and relative frequency of circulating variants in the territory (background bars proportional to the maximum rates, ranging from 0 to 100% relative frequency). The panel scales (y-axis) are independent and not proportional to each other. The figure presents the incidence and mortality rates of Influenza-like Illness (‘ILI’, mild and moderate cases) and Severe Acute Respiratory Syndrome (SARS) due to COVID-19 in Brazil between 01/01/2020 and 12/31/2024 (5 years), reporting respectively: incidence (A) and mortality (B) rates of mild and moderate cases, followed by incidence (C) and mortality (D) rates of severe cases.

Genomic surveillance data in Brazil were made available with limited sampling, providing the temporal distribution of circulating viral variants across the country and regions between 2020 and 2024. The absolute and relative frequencies of the variants among the total sequenced samples were analysed (Figure 2-A), allowing for a comparative assessment of lineage dynamics over the epidemiological weeks. An increasing concern with the surveillance was evident from the rise in sequencing efforts. However, as incidence and mortality rates declined after 2022, the number of sequencings also dropped sharply. In 2024, several circulating variants were recorded without specific lineage identification (categorized as ‘Others’), except for the sub-lineage JN.1 (KP.3*, XEC). No major differences were observed between regions regarding the relative frequencies of circulating variants (Figure 2-B).

**Figure 2.**
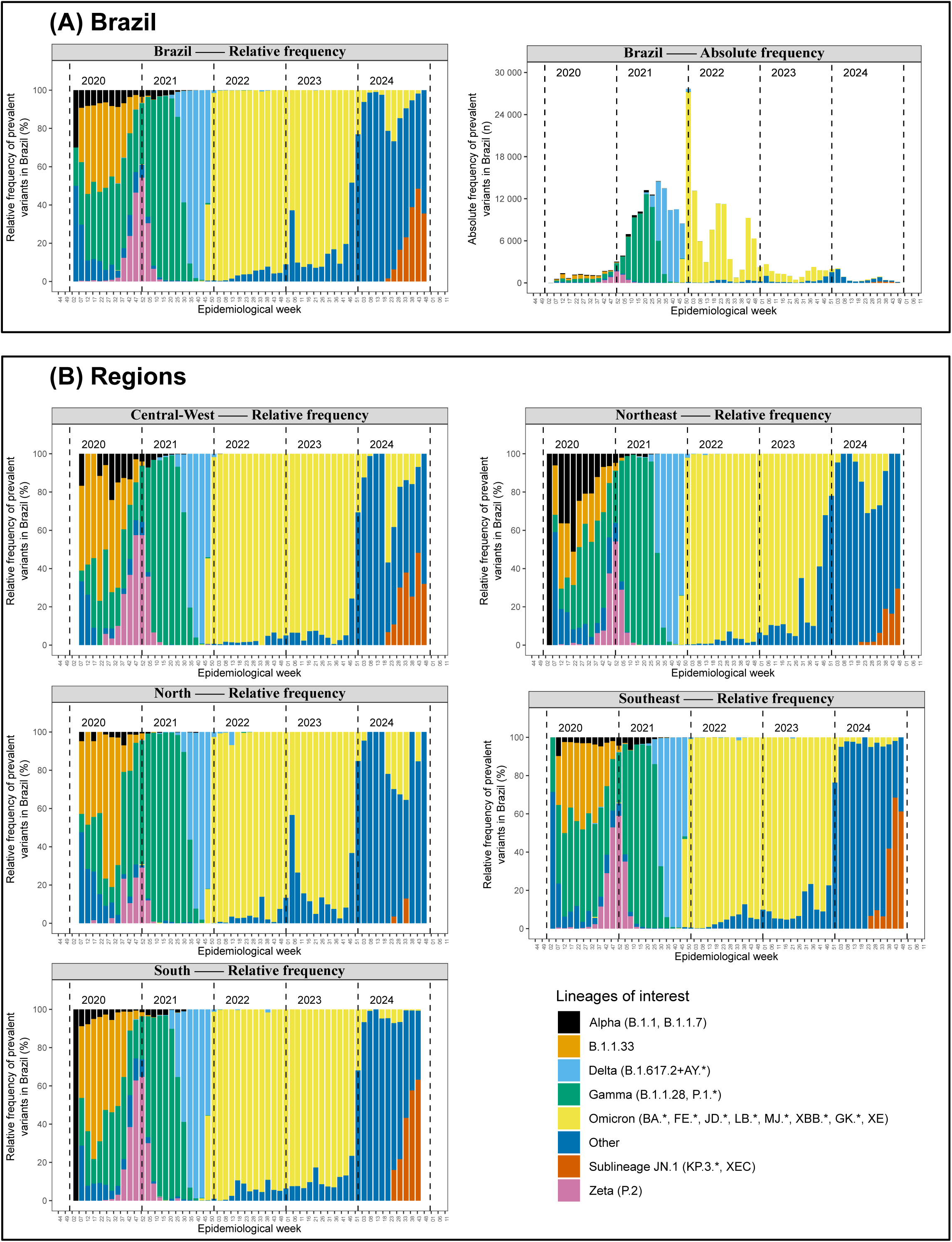
Relative (left) and absolute (right) frequency of circulating viral variants in Brazil (A) and its Regions (B) between 2020 and 2024. A large number of samples were collected between 2021 and 2022 (A - Absolute frequency), followed by a decline. No substantial differences are observed in the relative frequencies when comparing Brazil to its regions, or between the regions themselves (A-B). Lineages are categorized by distinct colours, as indicated in the legend.

The first pandemic wave was attributed to the variant of interest (VOI) B.1.1.33 and to the variants of concern (VOCs) Alpha (B.1.1, B.1.7). The second wave was associated with the prevalence of VOCs Delta (B.1.617.2+AY*) and Gamma (B.1.1.28, P.1*). The third wave was primarily driven by the circulation of the VOC Omicron (BA., FE., JD., LB., MJ., XBB., GK., XE), which became the predominant variant from the 45^th^ epidemiological week of 2021 until the beginning of 2024 (8^th^ week). Omicron also exhibited three distinct incidence peaks, each with progressively reduced amplitude, coinciding with the considerable progress of vaccination campaigns, particularly the administration of first booster doses (Figures 1-2; Figures S2-S3). After the reduction in these rates, unidentified variants (classified as “Others”) began to emerge. From the second quarter of 2024 onward, the rise of the VOI JN.1 (KP.3*, XEC) was observed, accompanied by new fluctuations in incidence and mortality rates. By the end of the fifth year, JN.1 sublineage variants had become predominant in both the Americas and Europe, where updated vaccines targeting these variants were already available^28^.

During the analysed period, 1,171,818 PCR-confirmed COVID-19 cases progressed to Severe Acute Respiratory Syndrome (SARS), of which 777,672 resulted in recovery and 394,129 in death, corresponding to a case fatality rate (CFR) of 33.63%. This rate varied significantly across all sociodemographic, socioeconomic, and health variables (χ², p<0.05) (Table 1). These variables - a total of 78 - were also fitted in univariate Generalized Linear Models (GLMs) (Table S1), and only those with p-value < 0.05 and odds ratio (OR) > 1.0 were retained for further analysis (Figure 3).

**Figure 3.**
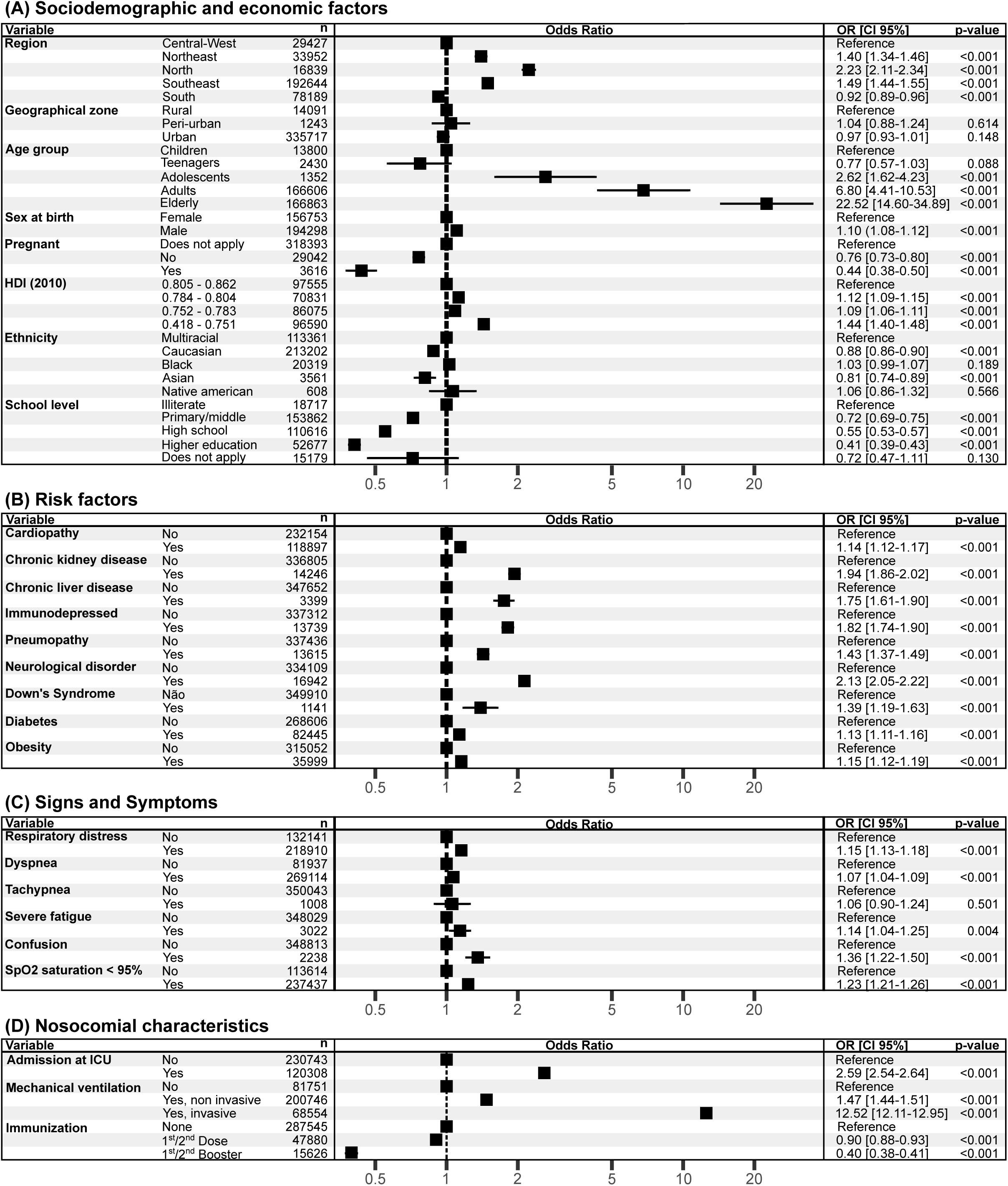
Forest plot presenting the Odds Ratios (OR) estimated from the binomial Generalized Linear Model, indicating the proportional association between independent variables and the occurrence of death for sociodemographic and economic factors (A), risk factors (B), signs and symptoms (C), and nosocomial factors (D). Each point represents the OR with 95% confidence intervals, and p-values < 0.05 indicate statistical significance. The X-axis displays the OR, with ‘1.0’ representing the reference level (no proportional effect relative to the reference group). OR values > 1.0 indicate a higher likelihood of death or aggravating factor, while OR < 1.0 indicate lower likelihood or a protective factor.

**Table 1.** Demographic characteristics of the population with PCR-confirmed cases of SARS due to COVID-19 in Brazil, from 01/01/2020 to 12/31/2024 (5 years) and for each year. The χ² test indicated significant differences between cure and death proportions across all strata (χ², p < 0.001). Case fatality rates (CFR%) were compared using the paired Z-Test (p < 0.05). Significance letters indicate statistically significant differences between groups; identical letters represent groupings with no significant difference. Proportions are presented in descending order (from highest to lowest).

|  |  | 01/01/2020 - 31/12/2024 (5 years) |  |  |  | 2020 |  |  |  | 2021 |  |  |  | 2022 |  |  |  | 2023 |  |  |  | 2024 |  |  |  |
| --- | --- | --- | --- | --- | --- | --- | --- | --- | --- | --- | --- | --- | --- | --- | --- | --- | --- | --- | --- | --- | --- | --- | --- | --- | --- |
| (n=1,171,801) | Factor level | Recovered | Deceased | CFR (%) | $\chi^2$ | Recovered | Deceased | CFR (%) | $\chi^2$ | Recovered | Deceased | CFR (%) | $\chi^2$ | Recovered | Deceased | CFR (%) | $\chi^2$ | Recovered | Deceased | CFR (%) | $\chi^2$ | Recovered | Deceased | CFR (%) | $\chi^2$ |
|  |  | 777,672 | 394,129 | 33.63 |  | 273,699 | 144,464 | 34.55 |  | 410,239 | 217,339 | 34.63 |  | 66,899 | 26,184 | 28.13 |  | 16,122 | 3,730 | 18.79 |  | 10,713 | 2,412 | 18.38 |  |
| Region (%) | Central-West | 82,488 (10.6) | 33,977 ( 8.6) | 29.17e | <0.001 | 29,961 ( 10.9) | 11,527 ( 8.0) | 27.78e | <0.001 | 42,624 (10.4) | 19,793 ( 9.1) | 31.71e | <0.001 | 6,765 (10.1) | 2,078 ( 7.9) | 23.5c | <0.001 | 1,942 (12.0) | 326 ( 8.7) | 14.37c | <0.001 | 1,196 (11.2) | 253 ( 10.5) | 17.46a | 0.034 |
|  | Northeast | 91,851 (11.8) | 70,172 (17.8) | 43.31b |  | 34,156 ( 12.5) | 31,754 ( 22.0) | 48.18b |  | 48,036 (11.7) | 33,383 (15.4) | 41b |  | 7,105 (10.6) | 4,310 (16.5) | 37.75a |  | 1,551 ( 9.6) | 461 (12.4) | 22.91a |  | 1,003 ( 9.4) | 264 ( 10.9) | 20.84a |  |
|  | North | 23,308 ( 3.0) | 20,938 ( 5.3) | 47.32a |  | 9,508 ( 3.5) | 9,616 ( 6.7) | 50.28a |  | 11,207 ( 2.7) | 10,045 ( 4.6) | 47.26a |  | 1,686 ( 2.5) | 1,010 ( 3.9) | 37.46a |  | 445 ( 2.8) | 149 ( 4.0) | 25.08a |  | 462 ( 4.3) | 118 ( 4.9) | 20.34a |  |
|  | Southeast | 433,662 (55.8) | 202,142 (51.3) | 31.79c |  | 154,643 ( 56.5) | 72,651 ( 50.3) | 31.96c |  | 229,084 (55.8) | 113,102 (52.0) | 33.05d |  | 36,184 (54.1) | 13,400 (51.2) | 27.02b |  | 8,419 (52.2) | 1,847 (49.5) | 17.99b |  | 5,332 (49.8) | 1,142 ( 47.3) | 17.64a |  |
|  | South | 14,6363 (18.8) | 66,900 (17.0) | 31.37d |  | 45,431 ( 16.6) | 18,916 ( 13.1) | 29.4d |  | 79,288 (19.3) | 41,016 (18.9) | 34.09c |  | 15,159 (22.7) | 5,386 (20.6) | 26.22b |  | 3,765 (23.4) | 947 (25.4) | 20.1a |  | 2,720 (25.4) | 635 ( 26.3) | 18.93a |  |
| Geographical Zone (%) | Urban | 668,843 (96.7) | 332,589 (95.6) | 33.21b | <0.001 | 236,978 ( 97.2) | 122,081 ( 95.7) | 34c | <0.001 | 352,255 (96.6) | 183,166 (95.6) | 34.21b | <0.001 | 56,589 (95.9) | 22,066 (95.3) | 28.05b | <0.001 | 13,754 (95.3) | 3,185 (94.0) | 18.8b | <0.001 | 9,267 (94.1) | 2,091 ( 93.3) | 18.41b | 0.001 |
|  | Peri-urban | 2,278 ( 0.3) | 1,192 ( 0.3) | 34.35b |  | 690 ( 0.3) | 471 ( 0.4) | 40.57b |  | 1,072 ( 0.3) | 613 ( 0.3) | 36.38b |  | 309 ( 0.5) | 84 ( 0.4) | 21.37c |  | 91 ( 0.6) | 12 ( 0.4) | 11.65b |  | 116 ( 1.2) | 12 ( 0.5) | 9.38c |  |
|  | Rural | 20,708 ( 3.0) | 14,186 ( 4.1) | 40.65a |  | 6,247 ( 2.6) | 5,002 ( 3.9) | 44.47a |  | 11,325 ( 3.1) | 7,840 ( 4.1) | 40.91a |  | 2,089 ( 3.5) | 1,014 ( 4.4) | 32.68a |  | 582 ( 4.0) | 192 ( 5.7) | 24.81a |  | 465 ( 4.7) | 138 ( 6.2) | 22.89a |  |
| Age group (%) | Children | 20,514 ( 2.6) | 1,093 ( 0.3) | 5.06d | <0.001 | 3,209 ( 1.2) | 328 ( 0.2) | 9.27c | <0.001 | 4,372 ( 1.1) | 337 ( 0.2) | 7.16d | <0.001 | 6,898 (10.3) | 253 ( 1.0) | 3.54c | <0.001 | 3,290 (20.4) | 97 ( 2.6) | 2.86c | <0.001 | 2,745 (25.6) | 78 ( 3.2) | 2.76c | <0.001 |
|  | Teenagers | 5,726 ( 0.7) | 291 ( 0.1) | 4.84d |  | 1,350 ( 0.5) | 90 ( 0.1) | 6.25d |  | 1,505 ( 0.4) | 83 ( 0.0) | 5.23d |  | 1,740 ( 2.6) | 85 ( 0.3) | 4.66c |  | 613 ( 3.8) | 19 ( 0.5) | 3.01c |  | 518 ( 4.8) | 14 ( 0.6) | 2.63c |  |
|  | Adolescents | 4,048 ( 0.5) | 417 ( 0.1) | 9.34c |  | 1,182 ( 0.4) | 156 ( 0.1) | 11.66c |  | 1,612 ( 0.4) | 190 ( 0.1) | 10.54c |  | 886 ( 1.3) | 51 ( 0.2) | 5.44c |  | 218 ( 1.4) | 9 ( 0.2) | 3.96c |  | 150 ( 1.4) | 11 ( 0.5) | 6.83c |  |
|  | Adults | 434,987 (55.9) | 111,999 (28.4) | 20.48b |  | 152,997 ( 55.9) | 31,755 ( 22.0) | 17.19b |  | 258,252 (63.0) | 75,253 (34.6) | 22.56b |  | 18,598 (27.8) | 4,037 (15.4) | 17.84b |  | 3,200 (19.8) | 558 (15.0) | 14.85b |  | 1,940 (18.1) | 396 ( 16.4) | 16.95b |  |
|  | Elderly | 312,397 (40.2) | 280,329 (71.1) | 47.29a |  | 114,961 ( 42.0) | 112,135 ( 77.6) | 49.38a |  | 144,98 (35.2) | 141,476 (65.1) | 49.47a |  | 38,777 (58.0) | 21,758 (83.1) | 35.94a |  | 8,801 (54.6) | 3,047 (81.7) | 25.72a |  | 5,360 (50.0) | 1,913 ( 79.3) | 26.3a |  |
| Sex at Birth (%) | Female | 351,932 (45.3) | 174,524 (44.3) | 33.15b | <0.001 | 122,997 ( 44.9) | 62,028 ( 42.9) | 33.52b | <0.001 | 180,993 (44.1) | 97,229 (44.7) | 34.95a | <0.001 | 34,031 (50.9) | 12,358 (47.2) | 26.64b | <0.001 | 8,370 (51.9) | 1,790 (48.0) | 17.62b | <0.001 | 5,541 (51.7) | 1,119 ( 46.4) | 16.8b | <0.001 |
|  | Male | 425,671 (54.7) | 219,566 (55.7) | 34.03a |  | 150,672 ( 55.1) | 82,419 ( 57.1) | 35.36a |  | 229,214 (55.9) | 120,091 (55.3) | 34.38b |  | 32,864 (49.1) | 13,823 (52.8) | 29.61a |  | 7,752 (48.1) | 1,940 (52.0) | 20.02a |  | 5,169 (48.3) | 1,293 ( 53.6) | 20.01a |  |
| Pregnant (%) | No | 78,342 (89.8) | 13,741 (94.7) | 14.92a | <0.001 | 28,093 ( 90.6) | 3,630 ( 95.3) | 11.44a | <0.001 | 44,015 (91.0) | 9,350 (94.1) | 17.52a | <0.001 | 4,796 (78.4) | 593 (97.9) | 11a | <0.001 | 915 (83.3) | 92 (96.8) | 9.14a | 0.001 | 523 (78.6) | 76 (100.0) | 14.53 | <0.001 |
|  | Yes | 8,925 (10.2) | 776 ( 5.3) | 8.0b |  | 2,899 ( 9.4) | 178 ( 4.7) | 5.78b |  | 4,379 ( 9.0) | 582 ( 5.9) | 11.73b |  | 1,321 (21.6) | 13 ( 2.1) | 0.97b |  | 184 (16.7) | 3 ( 3.2) | 1.6b |  | 142 (21.4) | 0 ( 0.0) | 0 |  |
| HDI 2010 (%) | 0.805 - 0.862 | 291,851 (38.0) | 106,963 (27.4) | 26.82d | <0.001 | 107,175 ( 39.6) | 38,346 ( 26.8) | 26.35c | <0.001 | 144,903 (35.8) | 58,463 (27.2) | 28.75d | <0.001 | 28,634 (43.4) | 8,364 (32.2) | 22.61d | <0.001 | 7,041 (44.5) | 1,126 (30.4) | 13.79c | <0.001 | 4,098 (38.8) | 664 ( 27.8) | 13.94c | <0.001 |
|  | 0.784 - 0.804 | 131,204 (17.1) | 67,216 (17.2) | 33.88c |  | 44,588 ( 16.5) | 24,601 ( 17.2) | 35.56b |  | 71,068 (17.6) | 36,687 (17.1) | 34.05c |  | 10,837 (16.4) | 4,713 (18.1) | 30.31c |  | 2,683 (17.0) | 751 (20.3) | 21.54b |  | 2,028 (19.2) | 464 ( 19.4) | 18.62b |  |
|  | 0.752 - 0.783 | 178,858 (23.3) | 97,960 (25.1) | 35.39b |  | 62,793 ( 23.2) | 35,101 ( 24.5) | 35.85b |  | 96,820 (23.9) | 54,916 (25.5) | 36.19b |  | 13,990 (21.2) | 6,515 (25.1) | 31.77b |  | 3,140 (19.8) | 862 (23.3) | 21.87b |  | 2,115 (20.0) | 566 ( 23.7) | 21.11ab |  |
|  | 0.418 - 0.751 | 165,921 (21.6) | 118,333 (30.3) | 41.63a |  | 56,040 ( 20.7) | 45,201 ( 31.6) | 44.65a |  | 92,101 (22.7) | 65,092 (30.3) | 41.41a |  | 12,488 (18.9) | 6,379 (24.6) | 33.81a |  | 2,963 (18.7) | 964 (26.0) | 24.55a |  | 2,329 (22.0) | 697 ( 29.2) | 23.03a |  |
| Ethnicity (%) | Caucasian | 352,229 (57.7) | 175,559 (53.1) | 33.26c | <0.001 | 116,055 ( 56.4) | 57,378 ( 49.4) | 33.08d | <0.001 | 188,701 (57.3) | 101,534 (54.5) | 34.98c | <0.001 | 33,802 (63.4) | 13,289 (59.1) | 28.22c | <0.001 | 8,232 (63.6) | 2,063 (62.6) | 20.04a | 0.12 | 5,439 (61.0) | 1,295 ( 60.2) | 19.23b | 0.003 |
|  | Multiracial | 218,777 (35.9) | 130,121 (39.4) | 37.29b |  | 74,429 ( 36.2) | 48,798 ( 42.0) | 39.6b |  | 120,592 (36.6) | 71,811 (38.5) | 37.32b |  | 16,595 (31.1) | 7,743 (34.4) | 31.81b |  | 4,081 (31.5) | 1,038 (31.5) | 20.28a |  | 3,080 (34.6) | 731 ( 34.0) | 19.18b |  |
|  | Black | 30,112 ( 4.9) | 20,302 ( 6.1) | 40.27a |  | 11,792 ( 5.7) | 8,133 ( 7.0) | 40.82a |  | 15,411 ( 4.7) | 10,771 ( 5.8) | 41.14a |  | 2,176 ( 4.1) | 1,156 ( 5.1) | 34.69a |  | 454 ( 3.5) | 140 ( 4.2) | 23.57a |  | 279 ( 3.1) | 102 ( 4.7) | 26.77a |  |
|  | Asian | 8,031 ( 1.3) | 3,927 ( 1.2) | 32.84c |  | 2,996 ( 1.5) | 1,614 ( 1.4) | 35.01cd |  | 4,125 ( 1.3) | 1,974 ( 1.1) | 32.37d |  | 687 ( 1.3) | 282 ( 1.3) | 29.1bc |  | 147 ( 1.1) | 39 ( 1.2) | 20.97a |  | 76 ( 0.9) | 18 ( 0.8) | 19.15ab |  |
|  | Native american | 1,068 ( 0.2) | 575 ( 0.2) | 35bc |  | 519 ( 0.3) | 316 ( 0.3) | 37.84abc |  | 392 ( 0.1) | 216 ( 0.1) | 35.53abcd |  | 84 ( 0.2) | 24 ( 0.1) | 22.22abc |  | 35 ( 0.3) | 15 ( 0.5) | 30a |  | 38 ( 0.4) | 4 ( 0.2) | 9.52ab |  |
| School level (%) | Illiterate | 9,596 ( 3.7) | 12,931 ( 8.8) | 57.4a | <0.001 | 3,446 ( 3.6) | 5,366 ( 10.2) | 60.89a | <0.001 | 4,435 ( 3.3) | 6,122 ( 7.4) | 57.98a | <0.001 | 1,272 ( 7.1) | 1,173 (12.4) | 47.95a | <0.001 | 278 ( 6.5) | 157 (10.5) | 36.09a | <0.001 | 165 ( 6.1) | 113 (11.9) | 40.65a | <0.001 |
|  | Primary/middle | 102,007 (39.7) | 79,312 (53.9) | 43.74b |  | 37,166 ( 38.4) | 29,006 ( 55.1) | 43.83b |  | 52,545 (38.9) | 43,389 (52.6) | 45.23b |  | 8,765 (49.0) | 5,450 (57.4) | 38.34b |  | 2,117 (49.5) | 911 (61.1) | 30.09a |  | 1,414 (52.7) | 556 ( 58.5) | 28.22b |  |
|  | High school | 95,342 (37.1) | 39,157 (26.6) | 29.11c |  | 35,617 ( 36.8) | 12,998 ( 24.7) | 26.74c |  | 52,934 (39.2) | 23,674 (28.7) | 30.9c |  | 4,902 (27.4) | 2,009 (21.2) | 29.07c |  | 1,137 (26.6) | 284 (19.0) | 19.99b |  | 752 (28.0) | 192 ( 20.2) | 20.36c |  |
|  | Higher education | 49,699 (19.4) | 15,695 (10.7) | 24d |  | 20,531 ( 21.2) | 5,241 ( 10.0) | 20.34d |  | 25,117 (18.6) | 9,369 (11.3) | 27.17d |  | 2,957 (16.5) | 856 ( 9.0) | 22.45d |  | 742 (17.4) | 139 ( 9.3) | 15.78b |  | 352 (13.1) | 90 ( 9.5) | 20.34c |  |

### Sociodemographic and Economic Factors

Regarding the Regions, the North (CFR: 47.32%; OR: 2.23 [2.11-2.34], p<0.05) and Northeast (CFR: 43.31%; OR: 1.40 [1.34-1.46], p<0.05) regions presented the worst scenarios, followed by the Southeast (CFR: 31.79%; OR: 1.49 [1.44-1.55], p<0.05) and South (CFR: 31.85%; OR: 0.92 [0.89-0.96], p<0.05), whereas the Central-West exhibited the lowest CFR at 29.61% (OR: Reference) (Table 1; Figure 3-A).

The vast majority of recorded cases were among individuals residing in urban areas (96.7% of recoveries and 95.6% of deaths). Although peri-urban and rural zones accounted for less than 5% of the total cases with recorded outcomes, an increase in CFR was observed with increasing distance from metropolitan centres (Table 1). Individuals from rural areas had the highest CFR (40.65%; OR: 1.03 [0.99-1.08]), followed by those from peri-urban zones (CFR: 34.35%; OR: 1.08 [0.92-1.27]), and finally urban residents (CFR: 33.21%; OR: *Reference*), although the differences between these groups were not statistically significant (Table 1; Figure 3-A).

Individuals aged between 0 and 59 months were classified as children, those between 5 and under 12 years as pre-adolescents, and those between 12 and under 18 years as adolescents. Individuals aged between 18 and under 60 years were classified as adults, and those aged 60 years and older as elderly. Regarding age classification, an increase in the case fatality rate (CFR) was observed with advancing age (Table 1). The lowest CFRs were found among children (CFR: 5.06%; OR: *Reference*) and pre-adolescents (CFR: 4.84%; OR: 0.77 [0.57-1.03], p>0.05), with no statistically significant difference between these two groups (Z-test and GLM, p>0.05). From adolescents, CFRs increased significantly (Z-test, p<0.05), showing a CFR of 9.34% (OR: 2.62 [1.62-4.23], p<0.05), followed by adults with a CFR of 20.48% (OR: 6.80 [4.41-10.53], p<0.05) and elderly individuals with a CFR of 47.29% (OR: 22.52 [14.60-34.89], p<0.05) (Table 1; Figure 3-A).

Male individuals exhibited a higher CFR (34.03%; OR: 1.10 [1.08-1.12], p<0.05) compared to females (CFR: 33.15%; OR: Reference). Among females, pregnant women aged between 15 and 45 years, regardless of whether gestational age was reported or not, had a lower CFR (8.0%; OR: 0.44 [0.38-0.50]) compared to non-pregnant women (CFR: 14.92%; OR: 0.76 [0.73-0.80]) (Z-test, p<0.05) (Table 1; Figure 3-A).

The 2010 Human Development Index (HDI) was adopted as a socioeconomic indicator, verified at the municipality level. Individuals were grouped according to HDI intervals from minimum to maximum values. A progressive increase in CFR was observed as the HDI decreased. Individuals residing in municipalities with HDI values between 0.418 and 0.751 had a CFR of 41.63% (OR: 1.44 [1.40-1.48], p<0.05); those between 0.752 and 0.783 had a CFR of 35.39% (OR: 1.09 [1.06-1.11], p<0.05); individuals from municipalities with HDI between 0.784 and 0.804 had a CFR of 33.88% (OR: 1.12 [1.09-1.15]); and those with HDI between 0.805 and 0.862 had the lowest CFR (26.82%; OR: *Reference*) (Table 1; Figure 3-A). Regarding ethnicity, Black individuals exhibited the highest CFR (40.27%; OR: 1.17 [1.12-1.21], p<0.05), followed by Multiracial individuals (CFR: 37.29%; OR: 1.14 [1.11-1.16], p<0.05). The lowest CFRs were observed among Asian (CFR: 32.84%; OR: 0.92 [0.84-1.01]), Native American People of Brazil (CFR: 35.00%; OR: 1.21 [0.98-1.50], p<0.05), and Caucasian individuals (CFR: 33.26%; OR: Reference), with no statistically significant differences among these latter groups (Z-test, p>0.05) (Table 1; Figure 3-A).

Regarding the Educational level, cases classified as ‘Not Applicable’ referred to individuals under 7 years of age. The highest CFR was observed among illiterate individuals (CFR: 57.4%; OR: *Reference*) and those with primary education (CFR: 43.74%; OR: 0.72 [0.69-0.75], p<0.05) (Z-test, p<0.05). A progressive reduction was observed across the subsequent levels: individuals with secondary education had a CFR of 29.11% (OR: 0.55 [0.53-0.57]), and those with higher education presented the lowest CFR (24.00%; OR: 0.41 [0.39-0.43]) (Z-test, p>0.05) (Table 1; Figure 3-A).

The risk factors retained in the model (Figure 3-B) were: cardiopathy (OR: 1.14 [1.12-1.17], p<0.05), chronic kidney disease (OR: 1.94 [1.86-2.02], p<0.05), chronic liver disease (OR: 1.75 [1.61-1.90], p<0.05), immunosuppression (OR: 1.82 [1.74-1.90], p<0.05), pneumopathy (OR: 1.43 [1.37-1.49], p<0.05), neurological disorder (OR: 2.13 [2.05-2.22], p<0.05), Down syndrome (OR: 1.39 [1.19-1.63], p<0.05), diabetes (OR: 1.13 [1.11-1.16], p<0.05) and obesity (OR: 1.15 [1.12-1.19], p<0.05).

The symptoms retained by the GLM model (Figure 3-C) were: respiratory distress (OR: 1.15 [1.13-1.18], p<0.05), dyspnea (OR: 1.07 [1.04-1.09], p<0.05), tachypnea (OR: 1.06 [0.90-1.24], p<0.05), severe fatigue (OR: 1.14 [1.04-1.25], p<0.05), confusion (OR: 1.36 [1.22-1.50]), and oxygen saturation (SpO₂) <95% (OR: 1.23 [1.21-1.26], p<0.05).

The nosocomial characteristics of the population analysed highlighted factors such as the presence of risk factors significantly influencing outcomes, with CFR increasing progressively with the number of pre-existing conditions, reaching 56.38% among individuals with three or more comorbidities (Figure 3-D; Table S2). Immunization also impacted clinical outcomes: individuals who were unvaccinated had a higher CFR (34.14%) compared to those who received booster dose (19.98%). The need for ICU support and invasive ventilation was strongly associated with death, with a CFR of 81.13% among patients requiring invasive ventilation (Figure 3-D; Table S2).

According to data from the National COVID-19 Vaccination Campaign, in addition to the primary vaccination series, additional, fractional, revaccination, and booster doses were administered. The COVID-19 SARS database includes information on ‘1^st^ Dose’ and ‘2^nd^ Dose’ as well as ‘1^st^ Booster’ and ‘2^nd^ Booster’ doses. Thus, this study focused on the primary vaccination series and booster doses.

Hospitalized individuals with COVID-19 SARS, when vaccinated, received doses of the following vaccines: ChAdOx1 nCoV-19 (AstraZeneca/Serum Institute), CoronaVac/Sinovac, BNT162b2 (Pfizer/BioNTech), Pfizer Baby, Pfizer Pediatric, Ad26.COV2.S (Janssen), VeroCell (Sinopharm), Covax (Covax Facility), and Gam-COVID-Vac (Sputnik V).

Approximately 85% of individuals who progressed to SARS had not been vaccinated (Table S2). The highest CFRs were observed among individuals who received only the 1^st^/2^nd^ doses (CFR: 35.65%; OR: 0.90 [0.88-0.93], p<0.05), although the multivariate model adjusted for various confounders indicated a 10% lower likelihood of death compared to the unvaccinated group (CFR: 34.14%; Reference). In contrast, those who received the 1^st^/2^nd^ booster showed a significant reduction in CFR, with up to a 60% lower likelihood of death compared to the unvaccinated group (CFR: 19.98%; OR: 0.40 [0.38-0.41], p<0.05) (Table S2; Figure 3-D).

The Kaplan-Meier survival curve analysis (Figure 4) revealed significant differences between the evaluated groups (log-rank test: χ² = 10,952; dF = 8; p<0.05). The curves illustrate the probability of survival over time for each group (Figure 4-A). Individuals who did not receive any vaccine dose (yellow line) exhibited the lowest probability of survival over time. Those who received the 1^st^/2^nd^ Dose (green line) showed improvement compared to the unvaccinated group but remained inferior to the group that received a booster. Individuals who received the 1^st^/2^nd^ Booster dose (blue line) displayed the highest probability of survival among all groups.

**Figure 4.**
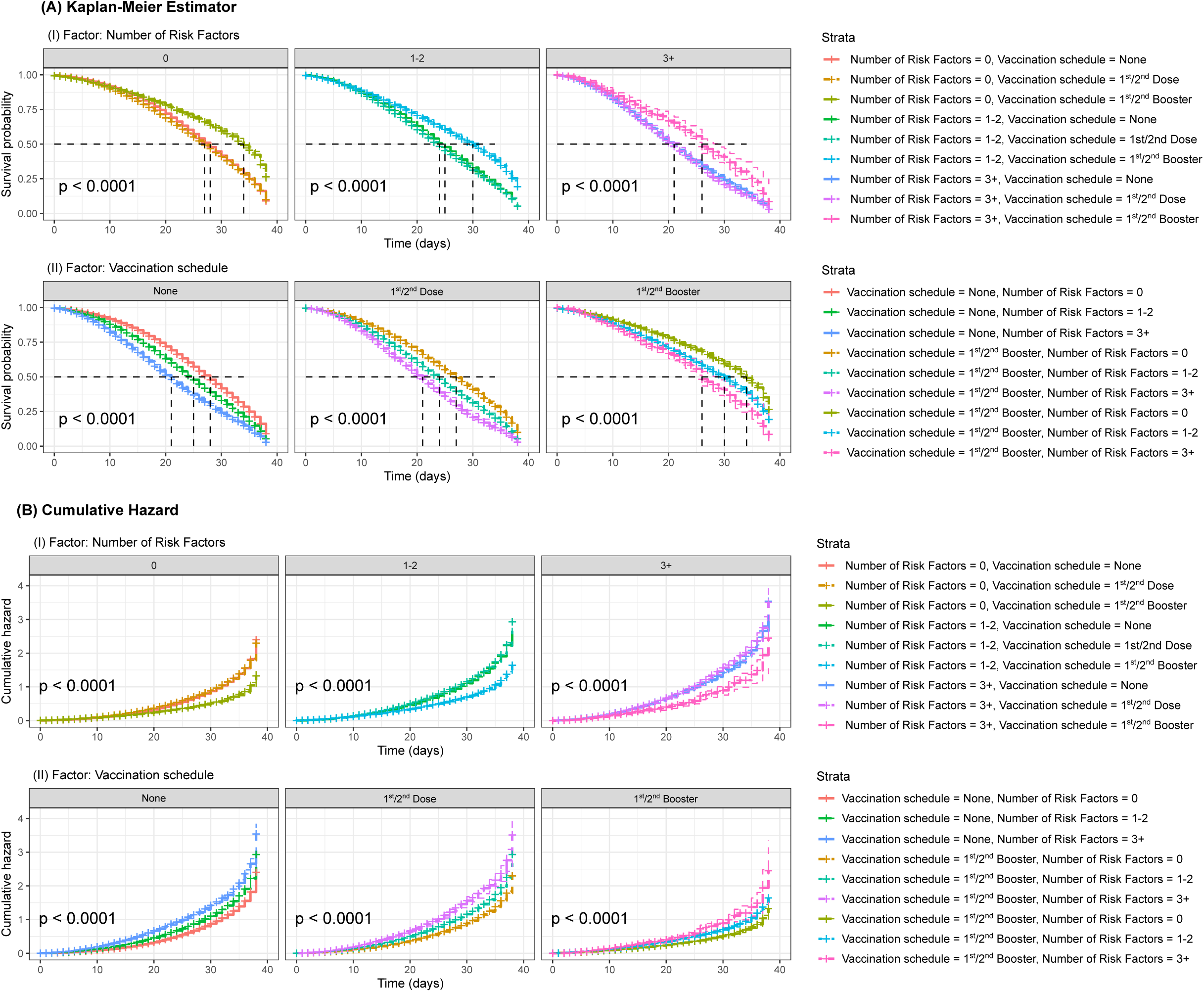
Kaplan-Meier survival curves stratified by number of comorbidities and vaccination regimen. In panel (A), the factor analysed is the number of comorbidities, with individuals categorized as: ‘0’ (no comorbidities/risk factors), ‘1-2’ (one or two comorbidities/risk factors), and ‘3+’ (three or more comorbidities/risk factors). In panel (B), the factor evaluated is the vaccination regimen, classified as: ‘None’ (no dose received), those immunized with one dose or with a complete primary schedule (‘1^st^/2^nd^ dose’), and those immunized with a 1^st^ or 2^nd^ booster (‘1^st^/2^nd^ Booster’). The curves show the probability of survival over time, with p-values indicating statistically significant differences (< 0.05) between groups.

Further, survival probability was analysed at specific time points (0, 10, 20, 30, and 40 days) based on the Kaplan-Meier model (Table S1). It was observed that individuals whose disease lasted for 30 days or longer showed a significant reduction in survival. Among unvaccinated individuals, the survival rate at 30 days was 39.0%, dropping to 6.4% at 40 days. For those who received the 1^st^/2^nd^ Dose, survival at 30 days was 37.0% and 6.0% at 40 days. In contrast, individuals who received 1^st^/2^nd^ Booster dose maintained a survival rate of 57.0% at 30 days, decreasing to 22.0% at 40 days.

## DISCUSSION

This study examines the dynamics of COVID-19 in Brazil between January 2020 and December 2024, considering mild and moderate cases reported by the Influenza-like Illness (ILI) surveillance system and severe cases requiring hospitalization due to Severe Acute Respiratory Syndrome (SARS). The research analyses interconnected factors such as the emergence of new variants, the vaccination schedule, the circulating variants across Brazilian territory, and the demographic and clinical characteristics of the susceptible population.

Brazil, with its continental dimensions, is marked by great diversity, reflected in its population. The different regions present unique demographic, social, economic, political and cultural characteristics. This heterogeneity, combined with the availability of public data, makes the country a valuable case for studying epidemiological dynamics, with results that can be extrapolated to global contexts.

### Limitations and Challenges for Epidemiological Surveillance in Brazil

One of the main challenges encountered in this study was the quality of the available data. While the public release of health information is a valuable initiative, significant gaps in data completeness were identified. Issues related to data quality were already present before the pandemic and have been documented across other national databases^29–32^. In addition to missing demographic variables, important clinical and epidemiological information was often absent. Another critical limitation was the delay in data reporting, which compromised the accuracy of compartmental epidemiological models and made real-time crisis management more difficult.

Furthermore, after the official end of the pandemic emergency phase, there was a marked decline in the number of samples collected for genomic surveillance.

### Emergence of Variants, Epidemiological Trends, and Regional Dynamics in Brazil

The SARS-CoV-2 variants rapidly acquired mutations that resulted in apomorphies enhancing their transmissibility and lethality, contributing to multiple outbreaks and culminating in public health crises such as those observed in Manaus^33,34^.

The Alpha variant carried mutations in the Spike protein, notably E484K, which enabled immune escape by reducing the effectiveness of vaccines and monoclonal antibody therapies^35–37^, as well as increasing both transmissibility^38^ and lethality. Additionally, the N501Y mutation was associated with enhanced binding to the ACE2 receptor and further immune escape^39^.

The Gamma variant, in addition to harbouring the E484K mutation, exhibited the D614G mutation, which increased replication efficiency in the respiratory tract and boosted transmissibility^40^ In the Delta variant, the T478K and L452R mutations enhanced ACE2 binding affinity and replication capacity^41^, while the P681R mutation was associated with increased pathogenicity^42^.

As the circulation of these variants declined, coinciding with reduced incidence and mortality rates and the expansion of vaccination efforts, the Omicron variant emerged. With up to 50 accumulated mutations^6^, Omicron became the predominant circulating variant by 2022 (Figures 1-2, Figures S2-S3). Although less pathogenic than Delta, Omicron exhibited alternative mechanisms for cellular entry and immune evasion, potentially contributing to a global reduction in vaccine effectiveness and demanding a third vaccine dose for appropriate protection^43^.

When analysing the circulating variant frequencies, the distribution dynamics were found to be remarkably similar across all regions of Brazil (Figure 2A-B). This suggests that the observed regional differences in pandemic are unlikely to be explained solely by variant circulation, which remained homogeneous nationwide. Therefore, other factors, such as, vaccination coverage, healthcare system capacity, socioeconomic conditions, and the local public health policies likely played a more prominent role in determining the epidemiological outcomes.

### Sociodemographic Profiles and Health Inequalities in the Brazilian COVID-19 Pandemic

Regarding the Brazilian regions, the North and Northeast exhibited the highest CFRs (Table 1), highlighting a scenario marked by limited access to and lower quality of healthcare services. In contrast, these regions reported the lowest adjusted incidence and mortality rates (Figures S2-S3). The vast geographic expanse of the North contributes to population dispersion, making access to hospitals more difficult and forcing residents of remote areas to travel to capital cities for specialized medical care^44,45^, as observed in the Amazonas state, where intensive care units are concentrated in the capital^46^. This issue is similarly seen in the Northeast region^47^ and in peri-urban and rural zones. Therefore, the problem in the North and Northeast was not primarily related to crisis management strategies (such as social distancing or lockdowns), but rather to the concentration of healthcare resources.

The elevated fatality rate among individuals aged 60 years and older is consistent with findings from other studies^48–50^, reflecting the effects of risk factors and the decline of immune function in this age group^51,52^. The increased case fatality rate among men was also observed, possibly attributable to differences in comorbidity profiles between sexes^49,53,54^.

In terms of ethnicity, it was observed that Black and Multiracial individuals experienced a higher number of deaths compared to Caucasians, in line with other studies reporting higher CFRs in these ethnic groups^55,56^. These differences may be associated with structural socioeconomic inequalities, reflecting historical disparities and intergenerational challenges in social mobility among racial groups. In Brazil, these inequalities are manifested in access to healthcare and across various social and economic dimensions, magnifying the impact Black and Multiracial populations^57,58^.

Furthermore, higher levels of education were associated with lower mortality rates, a finding consistent with studies indicating that education, along with other socioeconomic factors, is also a critical determinant of access to healthcare^56,59,60^. Low educational attainment is directly linked to higher rates of informal employment^61^, a condition that results in greater social vulnerability. The lack of labour protections for workers in the informal sector prevented many from adhering to social distancing measures during the pandemic, thereby increasing their exposure to SARS-CoV-2 and consequently raising incidence and mortality in these groups.

### From Comorbidities to Vaccination: Key Determinants of COVID-19 Severity

Since the beginning of the pandemic, studies have indicated that individuals with chronic diseases or risk factors exhibited more severe outcomes, allowing for the identification and prioritization of risk groups for medical interventions and vaccination campaigns. However, it remains challenging to establish a direct causal link between comorbidities and COVID-19 severity^62^.

During the 2003 outbreak of severe acute respiratory syndrome (SARS) caused by SARS-CoV-1, it was identified that ACE2 facilitated viral entry into cells. ACE2 is expressed in the bronchi, lungs, heart, kidneys, and gastrointestinal tract, aligning with the pathology observed in SARS-CoV-1 infections^63^. In 2020, it was confirmed that the new coronavirus, SARS-CoV-2, shared the same cellular entry mechanisms^64,65^. Transcriptomic and proteomic studies identified high ACE2 expression in tissues such as the intestines, kidneys, testes, and heart, moderate levels in the lungs, and elevated expression in type II pneumocytes^65–67^.

In addition to the lungs, SARS-CoV-2 likely targets other organs due to ACE2 expression, probably as a result of inflammatory processes and unresolved tissue injuries^68^. These processes can signal fibrosis and cardiac hypertrophy, vasoconstriction, endothelial dysfunction, and vascular inflammation, which may in turn trigger perivascular edema, systemic hypoxia, and disruptions in electrolyte balance, ultimately leading to multi-organ dysfunction^69,70^.

The findings reinforce the critical role of comorbidities and hospitalization severity in disease progression (Table S2). The negative impact of comorbidities was already anticipated, as patients with multiple pre-existing conditions often have a lower response capacity to treatment. Vaccination demonstrated a significant protective effect, reducing the CFR among vaccinated individuals, especially those who received booster doses. Prolonged hospitalization time among fatal cases suggests more severe disease evolution, possibly due to secondary complications. Higher CFR among patients requiring invasive ventilation reflects the severity of respiratory failure in these cases. These results highlight the importance of preventive strategies and reinforce the role of vaccination in mitigating the most severe outcomes of COVID-19.

Vaccination in Brazil began with the approval of CoronaVac and ChAdOx1 nCoV-19. The efficacy rates reported for authorization for these vaccines were 50.9% and 70.42%, respectively^71,72^. A few months later, the Pfizer/BioNTech vaccine (Comirnaty) reported an efficacy of 82% after the first dose, and the Janssen vaccine showed 85.4% efficacy among seronegative individuals and 76.4% against severe disease^73,74^. Studies have shown that regardless of the vaccine platform, there was a significant increase in efficacy after the complete vaccination schedule^75–78^ (Table 2-3).

The survival curves demonstrate a significant benefit associated with booster doses, especially compared to those who received no vaccination (Figure 4). The 50% survival intercept point was considerably longer in the group that received a booster, suggesting an increased median survival time. Furthermore, among the most severe cases, starting at 30 days, the group that received a booster exhibited an 18 to 20 percentage point higher survival rate compared to other groups, and approximately four times higher survival at 40 days (Table S1). This analysis reinforces the effectiveness of booster doses to mitigate or prevent COVID-19 SARS cases.

## OPERATIONAL AND PUBLIC POLICY RECOMMENDATIONS

Based on the findings of this study, it becomes evident that structural and operational advancements are needed within the Brazilian healthcare system, especially regarding preparedness for future public health emergencies across different domains.

One of the main proposals involves strengthening policies aimed at expanding hospital services into rural and underserved areas, with an emphasis on increasing the number of intensive care unit beds and ensuring the equitable distribution of healthcare professionals in remote and peri-urban regions, historically lacking adequate coverage. Beyond physical infrastructure, the continuous training and support of health professionals is also essential, with incentives to retain them in hard-to-reach locations.

Another key area relates to the modernization of health information systems. The results revealed significant delays in data entry and a high rate of missing values for essential variables, which undermined real-time analysis and coordinated responses to the crisis. It is therefore recommended to continuously improve mechanisms for data collection, integration, and interoperability, through investments in digital technologies and ongoing training for the personnel responsible for data registration. This strategy should be aligned with the promotion of partnerships among federal entities, universities, and research centres to develop public platforms for automated analysis and evidence-based decision-making.

Additionally, the creation of a permanent federal fund for health emergencies is proposed, with the capacity for immediate activation in the event of a calamity. This fund should prioritize support for the most vulnerable populations, such as informal workers and individuals without access to labour protections, enabling effective compliance with social distancing measures without jeopardizing families’ food and economic security. Registration of these populations could be integrated into the CadÚnico system, with regular updates and automated activation of emergency aid. This measure aims to reduce the population’s sense of insecurity, considering that social distancing rules were established on March 11^79^, yet emergency financial aid was only approved on March 30^80^ and became available for withdrawal on April 9, nearly a month later^81^, which was unfeasible for informal workers who rely on subsistence income.

Genomic surveillance, in turn, must be treated as a central component of the response to future pandemics. The heterogeneity and discontinuity of genomic monitoring in Brazil, as evidenced by the data presented, reveal the vulnerability of the country’s capacity to track and respond to variants of concern.

Finally, it is essential to develop a public sector communication plan tailored to the sociocultural contexts of different Brazilian regions. Public health campaigns on vaccination and preventive measures must be conducted in accessible language. Such measures are crucial to reducing vaccine hesitancy, combating misinformation, and increasing public engagement in health initiatives.

## CONCLUSIONS

The results highlight the complexity and regional inequalities in the response to the COVID-19 pandemic in Brazil, revealing significant variations in case fatality rates and public health responses. Three major epidemic waves were observed, with the second being the most impactful across all regions. The circulation of variants such as Gamma and Omicron played a crucial role in the pandemic’s dynamics, influencing incidence, mortality, and regional patterns^82^, closely related to crisis management strategies involving social distancing and the distribution of hospital resources.

Incidence and mortality varied significantly between regions. The most vulnerable groups included the elderly, Black and Multiracial individuals, and people with low levels of education. Residents of rural areas exhibited higher lethality compared to those in urban areas, reflecting disparities in access to healthcare services.

Vaccination had a positive impact on reducing case fatality rates, particularly following the administration of booster doses. Unvaccinated individuals exhibited significantly higher mortality rates. Despite advances in the immunization schedule vaccine adherence remained uneven, and efficacy was limited in contexts of high variant circulation.

The progression of the pandemic in Brazil was deeply influenced by social and structural inequalities, disproportionately affecting vulnerable populations. Genomic surveillance, policies supporting social distancing, and vaccination were fundamental in mitigating the effects of the pandemic but faced significant limitations due to delayed implementation and unequal access. The findings reinforce the urgent need to strengthen healthcare systems, raise the concern for the constant training of employees responsible to collect and record secondary health data, expand access to vaccination, and promote continuous monitoring strategies to enhance preparedness for future public health emergencies.

## Data Availability

All bash and R script/codes will be released on GitHub's Corresponding Author upon publishing of the material.

## ACKNOWLEDGEMENTS

The authors thank the Brazilian Government and the Brazilian Ministry of Health for making public secondary data available and acknowledge all individuals directly and indirectly involved in the efforts to manage the COVID-19 crisis in the country.

Silvio Alencar Cândido-Sobrinho gratefully acknowledges the Coordination for the Improvement of Higher Education Personnel (CAPES/MEC/BRAZIL) for the study grant (grant number: 88887.479534/2020-00) - Finance Code 001.

## DATA SHARING

Bash and R scripts used in this research will be made available at the Corresponding Author’s GitHub upon publishing in a journal.

---

**Figure S1.**
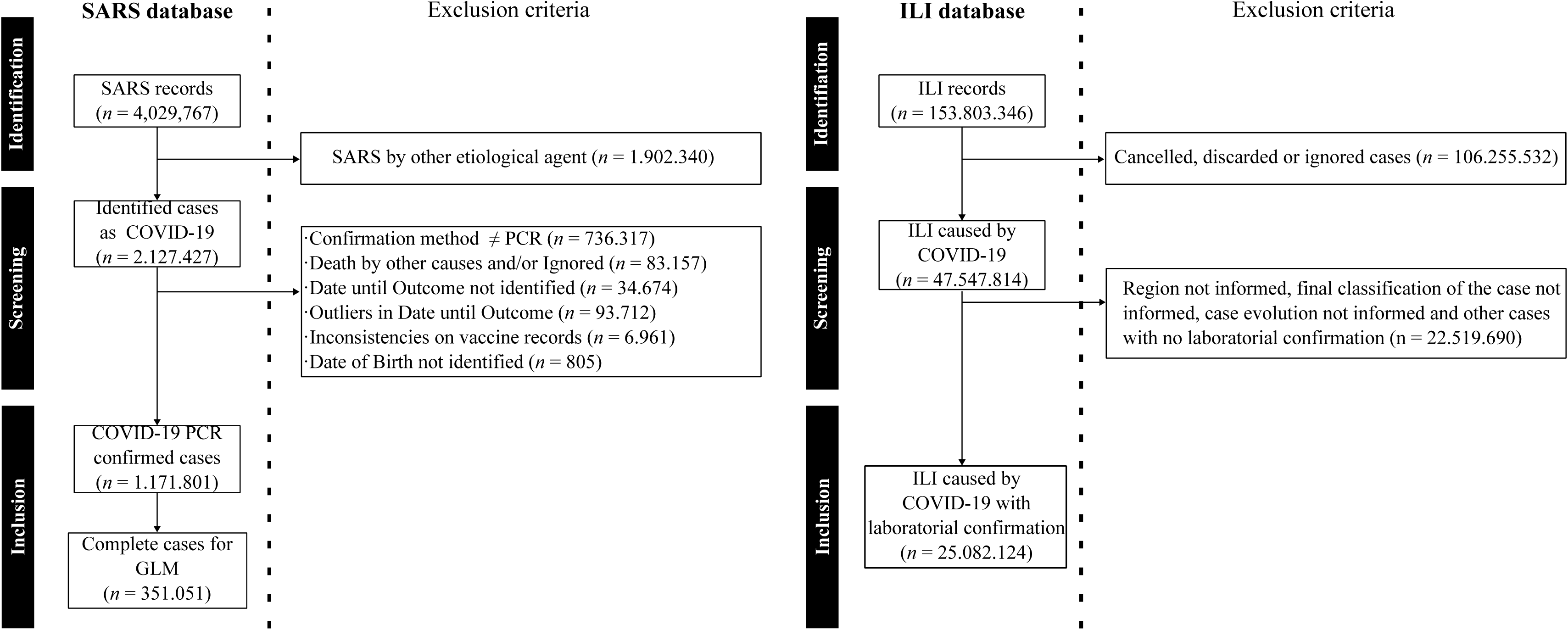
Eligibility criteria for patients with mild, moderate, and severe cases reported in Brazil from 01/01/2020 to 12/31/2024.For the ILI database, cases with ignored, discarded, or cancelled classifications were excluded, as well as those without a reported region and those classified by a non-laboratory method. For the SARS database, cases with other etiological agents were excluded, along with those lacking birth or closure dates, those with ignored outcomes or deaths due to other causes, those with discrepant closure dates, and those classified with an etiological agent identification method other than PCR.

**Figure S2.**
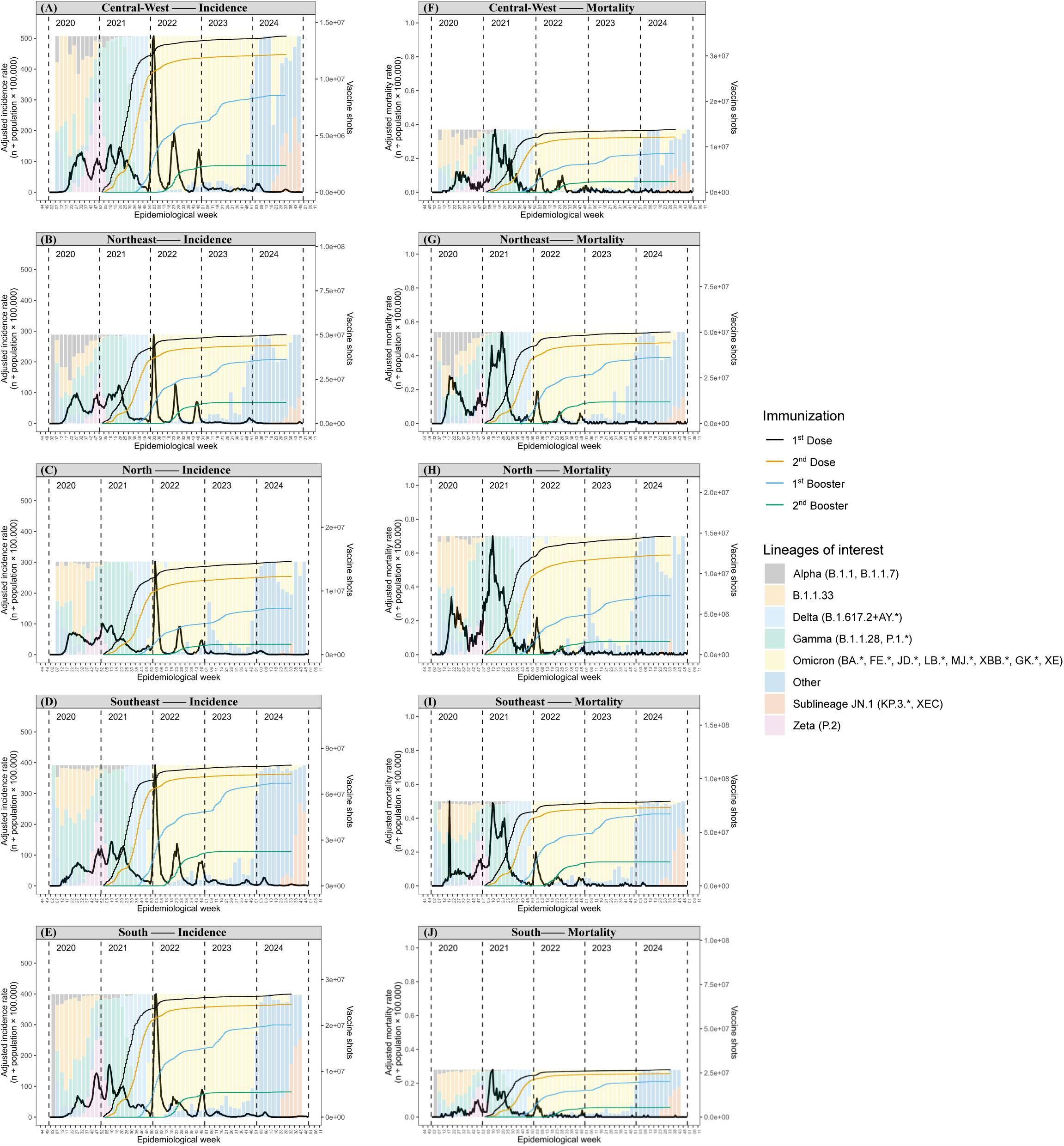
**A**djusted incidence and mortality rates of ILI (Influenza-like Illness) cases (left y-axis) by epidemiological week (x-axis), relative frequency of circulating variants (background bars scaled to the maximum values between 0 and 100% relative frequency), and vaccination trends observed across different regions of Brazil between 01/01/2020 and 12/31/2024 (5 years). Incidence rates were irregular (A-E), and Omicron was the most prevalent variant. Few deaths were recorded in the ILI dataset, resulting in case fatality rates below 1 per 100,000 inhabitants (F-J).

**Figure S3.**
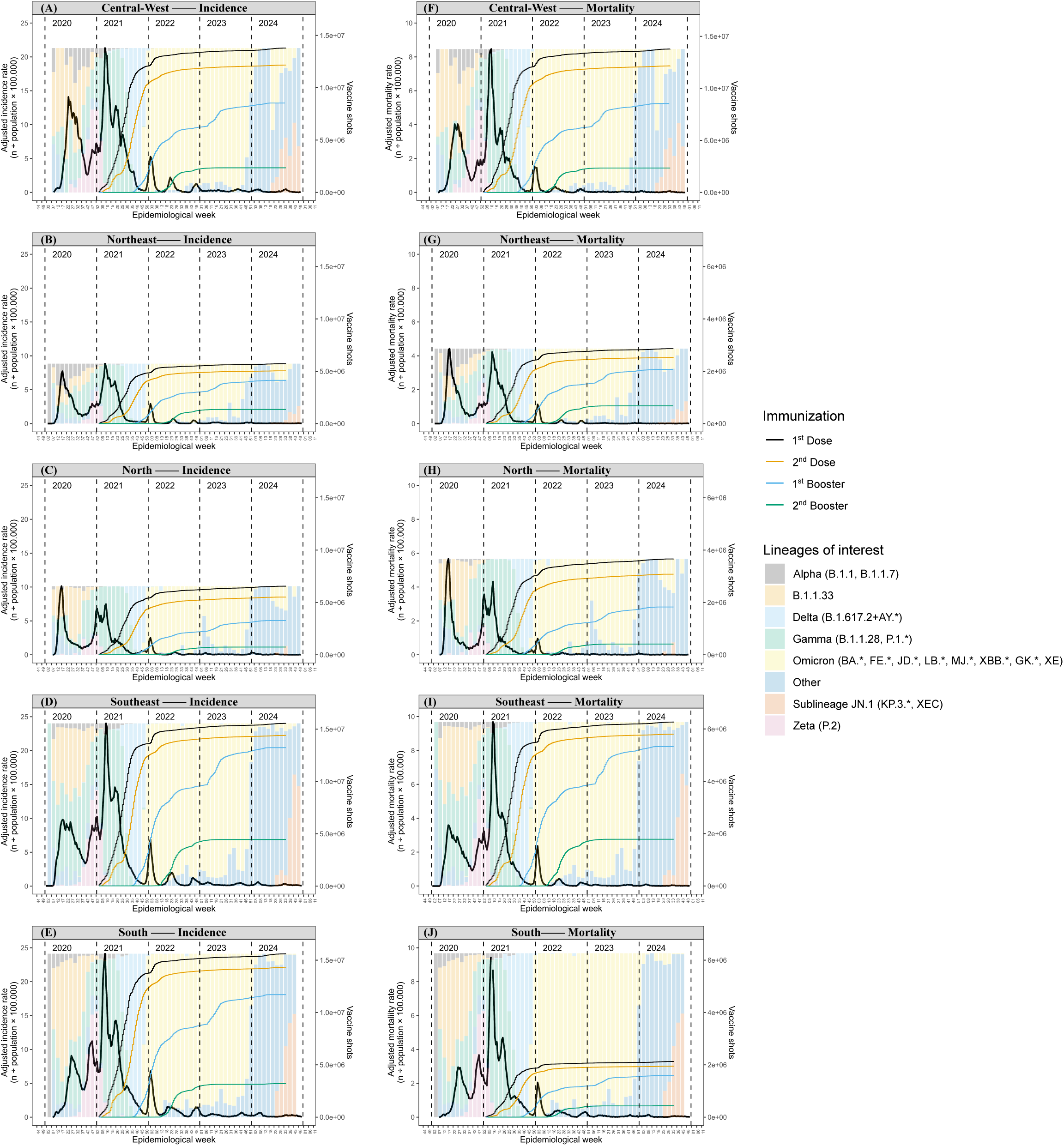
Adjusted incidence and mortality rates of SARS (Severe Acute Respiratory Syndrome) cases (left y-axis) by epidemiological week (x-axis), relative frequency of circulating variants (background bars scaled to the maximum values between 0 and 100% relative frequency), and vaccination trends observed across different regions of Brazil between 01/01/2020 and 12/31/2024 (5 years). Three distinct waves were observed during the Public Health Emergency period in the North and Northeast regions (B-C), while other regions showed up to five waves (Central-West, Southeast - A, D) or more (South - E). The highest mortality rates coincided with the widespread circulation of the Gamma variant across all regions, highlighting it as the most impactful variant on incidence and mortality during 2021 (F-J).

**Table S1.**
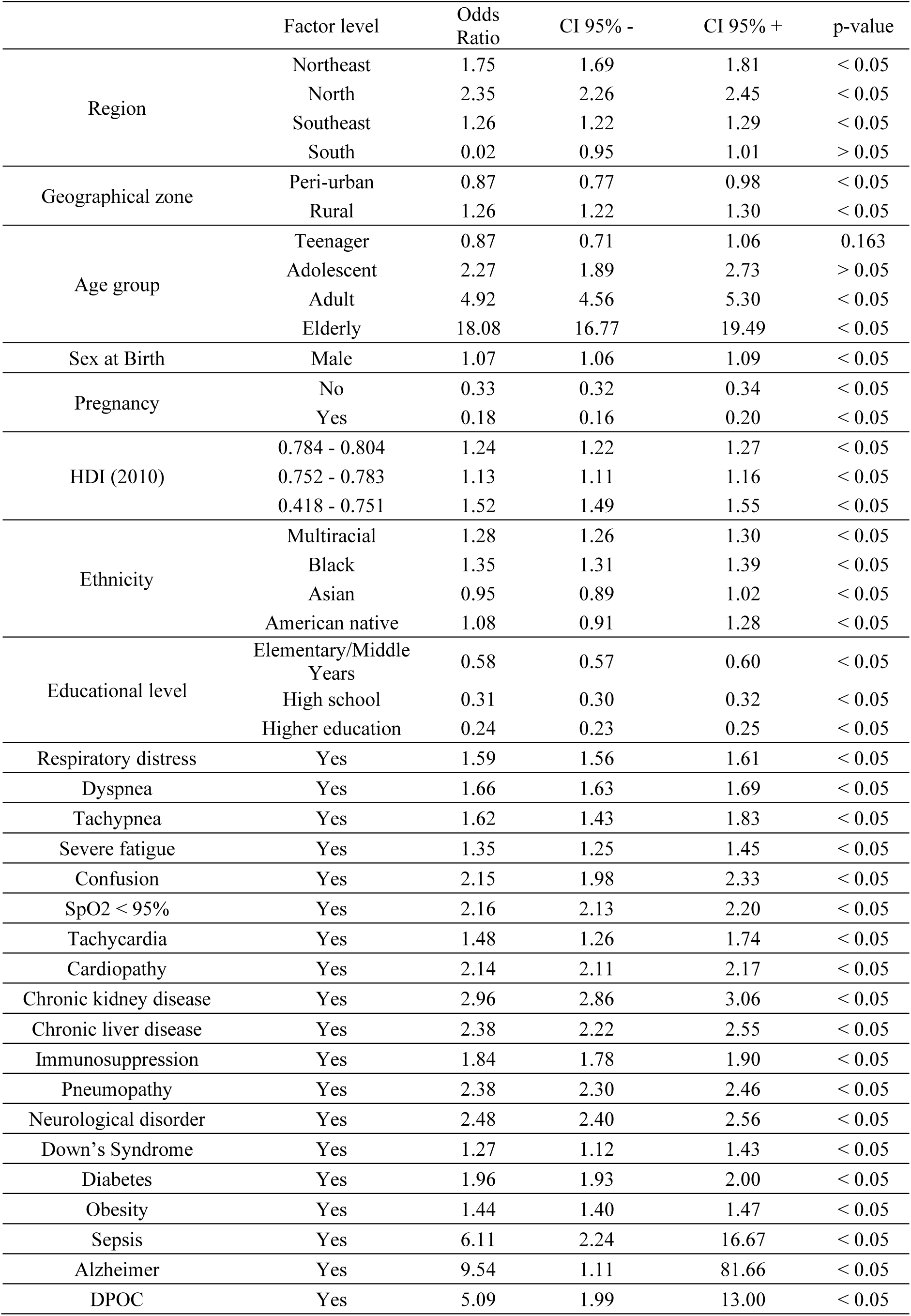
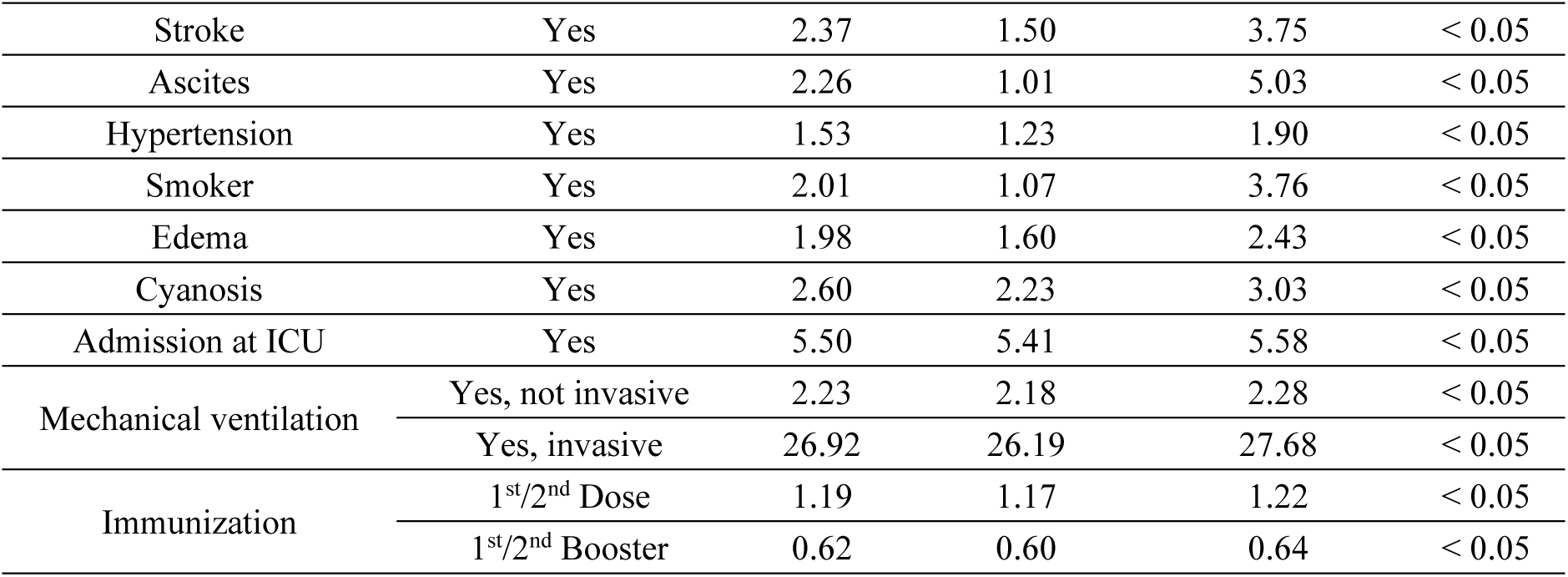
Odds Ratios and corresponding confidence intervals from univariate generalized linear models with p-values < 0.05 and OR > 1.0.

**Table S2.** Nosocomial characteristics of the population with confirmed cases of SARS by COVID-19 through PCR methodology in Brazil by year, between 01/01/2020 and 12/31/2024 (5 years). The χ² test indicated significant differences between cure and death proportions across all strata (χ², p < 0.001). Case fatality rates (CFR%) were compared using the paired Z-test (p < 0.05). Significance letters indicate statistically significant differences between groups, where similar letters represent groupings, and proportions are organized in descending order (from highest to lowest proportion).

| (n=1,171,801) | Factor level | Recovered | Deceased | CFR (%) | $\chi^2$ Test |
| --- | --- | --- | --- | --- | --- |
|  |  | 777,672 | 394,129 | 33.63 |  |
| Number of comorbidities (%) | 0 | 438,780 (56.4) | 163,762 (41.6) | 27.18c | <0.001 |
|  | 1-2 | 333,263 (42.9) | 223,091 (56.6) | 40.1b |  |
|  | 3+ | 5,629 ( 0.7) | 7,276 ( 1.8) | 56.38a |  |
| Immunization (%) | None | 637,970 (82.0) | 330,637 (83.9) | 34.14b | <0.001 |
|  | 1<sup>st</sup>/2<sup>nd</sup> Dose | 94,039 (12.1) | 52,092 (13.2) | 35.65a |  |
|  | 1<sup>st</sup>/2<sup>nd</sup> Booster | 45,663 ( 5.9) | 11,400 ( 2.9) | 19.98c |  |
| ICU (%) | No | 604,875 (77.8) | 171,478 (43.5) | 22.09b | <0.001 |
|  | Yes | 172,797 (22.2) | 222,651 (56.5) | 56.3a |  |
| Mechanical ventilation (%) | No | 196,463 (28.5) | 29,710 ( 8.8) | 13.14c | <0.001 |
|  | Yes, non-invasive | 454,070 (65.9) | 141,030 (41.8) | 23.7b |  |
|  | Yes, invasive | 38,729 ( 5.6) | 166,512 (49.4) | 81.13a |  |

**Table S3.** Survival probabilities estimated using the Kaplan-Meier model at time points 0, 10, 20, 30, and 40 days. The study population was stratified as: unvaccinated, vaccinated with the 1^st^ or 2^nd^ dose, or vaccinated with the 1^st^ or 2^nd^ booster.

| Vaccine Shots: None |  |  |  |  |  |  |
| --- | --- | --- | --- | --- | --- | --- |
| Time (days) | Numbers at Risk | Number of Events | Survival Probability | Standard-error | Lower CI95% | Upper CI95% |
| 0 | 970259 | 3829 | 0.996 | 6.36E-05 | 0.996 | 0.996 |
| 10 | 779779 | 88340 | 0.900 | 3.15E-04 | 0.899 | 0.900 |
| 20 | 287537 | 130590 | 0.679 | 6.03E-04 | 0.678 | 0.680 |
| 30 | 78458 | 77565 | 0.390 | 9.01E-04 | 0.388 | 0.392 |
| 40 | 4521 | 33065 | 0.064 | 1.05E-03 | 0.062 | 0.066 |
| Vaccine Shots: 1<sup>st</sup> or 2<sup>nd</sup> Dose |  |  |  |  |  |  |
| Time (days) | Numbers at Risk | Number of Events | Survival Probability | Standard-error | Lower CI95% | Upper CI95% |
| 0 | 142153 | 640 | 0.996 | 1.78E-04 | 0.995 | 0.996 |
| 10 | 108438 | 14717 | 0.884 | 8.87E-04 | 0.882 | 0.885 |
| 20 | 39754 | 20083 | 0.644 | 1.64E-03 | 0.641 | 0.648 |
| 30 | 11346 | 11217 | 0.368 | 2.28E-03 | 0.363 | 0.372 |
| 40 | 672 | 4784 | 0.059 | 2.59E-03 | 0.054 | 0.065 |
| Vaccine Shots: 1<sup>st</sup> or 2<sup>nd</sup> Booster |  |  |  |  |  |  |
| Time (days) | Numbers at Risk | Number of Events | Survival Probability | Standard-error | Lower CI95% | Upper CI95% |
| 0 | 46844 | 224 | 0.995 | 3.19E-04 | 0.995 | 0.996 |
| 10 | 29458 | 3626 | 0.904 | 1.49E-03 | 0.901 | 0.907 |
| 20 | 10357 | 3089 | 0.754 | 2.86E-03 | 0.749 | 0.760 |
| 30 | 3149 | 1537 | 0.569 | 4.84E-03 | 0.559 | 0.578 |
| 40 | 215 | 796 | 0.220 | 1.11E-02 | 0.199 | 0.243 |

